# A flexible multi-sensor device enabling handheld sensing of heart sounds by untrained users

**DOI:** 10.1101/2024.10.09.24315183

**Authors:** Andrew McDonald, Maximilian Nussbaumer, Nirmani Rathnayake, Richard Steeds, Anurag Agarwal

## Abstract

Heart valve disease has a large and growing burden, with a prognosis worse than many cancers. Screening with a traditional stethoscope is underutilised, often inaccurate even in skilled hands, and requires time-consuming, intimate examinations. Here, we present a handheld device to enable untrained users to record high-quality heart sounds without the need to undress. The device incorporates multiple high-sensitivity sensors embedded in a flexible substrate, placed at key chest locations by the user. To address challenges from localised heart sound vibrations and noise interference, we developed time-frequency signal quality algorithms that automatically select the best sensor in the device and reject recordings with insufficient diagnostic quality. A validation study demonstrates the device’s effectiveness across a diverse range of body types, with multiple sensors significantly increasing the likelihood of a successful recording. The device has the potential to enable accurate, accessible, low-cost heart disease screening.

## 1 Main

The stethoscope is a classic medical device for assessing bodily functions by listening to acoustic vibrations (auscultation). It is of particular importance for the first detection of heart murmurs indicating valvular heart disease (VHD), which is one of the most common causes of heart failure and has been described as the ‘next cardiac epidemic’ [1]. More than half of patients with significant VHD remain undiagnosed [2], and more than half of patients in Europe receive surgery too late, when intervention is less effective [3].

A key reason for this underdiagnosis is that accurate cardiac auscultation requires significant skill and can be a time-consuming and intimate process. A stethoscope exam requires a patient to undress and for the small chest piece to be placed carefully at key locations on the chest, including under the left breast [4]. Because of these factors, auscultation was only performed on 38% of UK patients with symptoms indicative of VHD [5], and female patients are less likely to receive a complete cardiac exam [6].

Even when a stethoscope exam is performed, the overall diagnostic accuracy of primary clinicians is poor [7]. General practitioners miss more than half of significant asymptomatic cases of VHD [8], and unnecessary referrals for innocent or absent murmurs are common [9].

In recent years, machine learning algorithms to automatically diagnose heart sound recordings have shown promising performance [10–13]. However, these algorithms still rely on a good quality signal recorded by a digital stethoscope [14]. Given the significant skill required in auscultation, digital recordings are often poor quality [14, 15]. This limits the reliable use of digital stethoscope screening to skilled clinicians in traditional primary care environments.

Designing a device with a large sensing area over the thorax increases the likelihood of capturing high-quality signals from the key auscultation sites, eliminating the need for skilled placement [16]. Previous studies have designed large spatial arrays of adhesive sensors [17, 18]. However, sticking sensors onto the skin is time-consuming, intrusive, and often painful. Other studies have proposed wearable sensor arrays that are pressed against the chest using straps or vests [16, 19–21]. However, designing a wearable device that can conform to a wide range of body shapes and breast sizes is challenging. Dressing a patient in a wearable ‘jacket’ may also be cumbersome and time-consuming, requiring intimate contact between the clinician and the patient. None of these devices have demonstrated formal usability studies that would be suitable for regulatory approval.

Crucially, most of these proposed devices require subjects to undress to apply the device on a bare chest [16–20]. This is a significant barrier to the widespread adoption of screening, particularly for female patients [22, 23]. With appropriate application force, piezoelectric sensors have been shown to obtain good-quality recordings through clothing [22]. Therefore, the need to undress may be overcome by a device design that does not rely on adhesives or wearables.

In this work, we present a proof-of-concept design and the first validation of a novel acoustic device that enables quick and easy self-collection of a patient’s heart sound data, removing the need for an intimate examination by an external clinician. Paired with machine learning algorithms [24], this device has the potential to enable widespread screening of VHD to be carried out by non-skilled operators in resource-strained primary care and community settings. This is made possible through (1) using multiple high-sensitivity piezoelectric sensors, optimally arranged in a flexible substrate, (2) keeping the device large enough to allow imprecise placement and small enough to fit in the palm of a hand, and (3) developing signal processing techniques to automatically assess the signal quality of recordings and provide feedback to the user.

## 2 Results

### 2.1 Mechanical device design

The device, shown in Fig. 1a, consists of six piezoelectric sensors arranged in a pentagonal pattern within a flexible 10.5 cm diameter substrate, ensuring easy handling for over 95% of users [25]. The sensors are spaced with a maximum separation of 5 cm, ensuring there are no sensing gaps large enough to miss localised heart sounds, thus providing comprehensive coverage of the sensing area around each auscultation site. Each sensor houses a PZT transducer (Fig. 1b), chosen for its insensitivity to airborne noise and reliability in vibration detection.

**Fig. 1.**
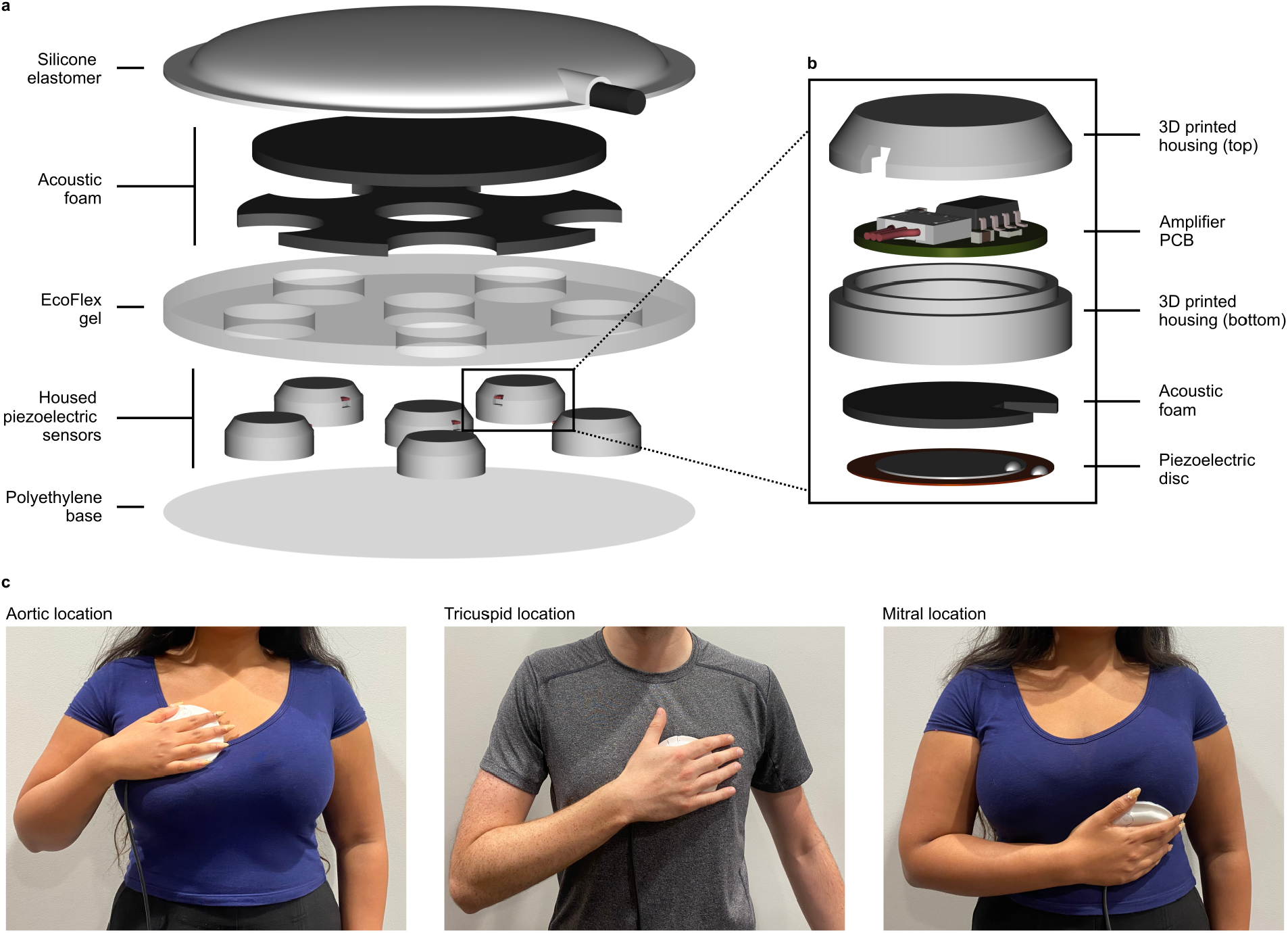
Design and example usage of sensor prototype. **a**, Exploded schematic of full sensor prototype. In this prototype, the housed sensors are connected to an external data acquisition system through flexible wires (not shown). A future design could be made wireless using a Bluetooth connection. **b**, Exploded view of one housed piezoelectric sensor. **c**, The prototype held by subjects at three key auscultation locations on the chest. The flexibility of the device allows it to conform to the curved surface of the chest, which is particularly important under the left breast at the mitral location.

The flexible substrate, made of silicone gel, maintains sensor positioning while minimising mechanical coupling. This ensures independent chest vibration measurements, with the device conforming to curved areas, especially around the left breast. The overall contact area is 87 cm^2^, with only 15 cm^2^ occupied by rigid components (see Extended Fig. E1).

Fig. 1c shows the application of the device at the key auscultation locations. Traditionally, four locations are used for cardiac auscultation. However, the pulmonic auscultation site is less critical for screening acquired valvular heart disease, as most VHD primarily affects the aortic and mitral valves. Consequently, the pulmonic location is often excluded in clinical studies focused on these more common left-sided valve pathologies [8]. Further design details and material properties are provided in Section 4.1.

### 2.2 Signal acquisition and benchmarking

Each sensor contains a 15 mm diameter PZT disc housed in a 3D-printed casing. The PZT material is sandwiched between a brass electrode, which forms the outer layer, and a smaller silver electrode. When the sensor is pressed against the chest, vibrations cause the brass electrode to bend, which in turn deforms the PZT material, generating an electric charge. This charge is converted into a voltage signal by an integrated charge amplifier, optimized with a 1 nF feedback capacitance and 4.5 V supply to maximise dynamic range. Low-frequency artefacts, such as hand tremors, are reduced by a 20 Hz high-pass filter, and the sensor is shielded to minimize electrical interference. The six sensors are sampled simultaneously at 5,120 Hz.

Figure 2 presents a comparison of our device sensor and the Littmann 3200 stethoscope, a standard reference for benchmarking, in response to identical excitation on a laboratory phantom. Both sensors’ outputs are referenced to their mean noisefloors in the 500–1,000 Hz range (detailed in section 4.2). The excitation level is tuned to give a fairly flat response for the Littmann 3200 at an average amplitude of 36 dB above its reference noise floor. Below 200 Hz, typical heart sounds are expected to exceed this amplitude. Under these conditions, both sensors produce signals well above their noisefloors.

**Fig. 2.**
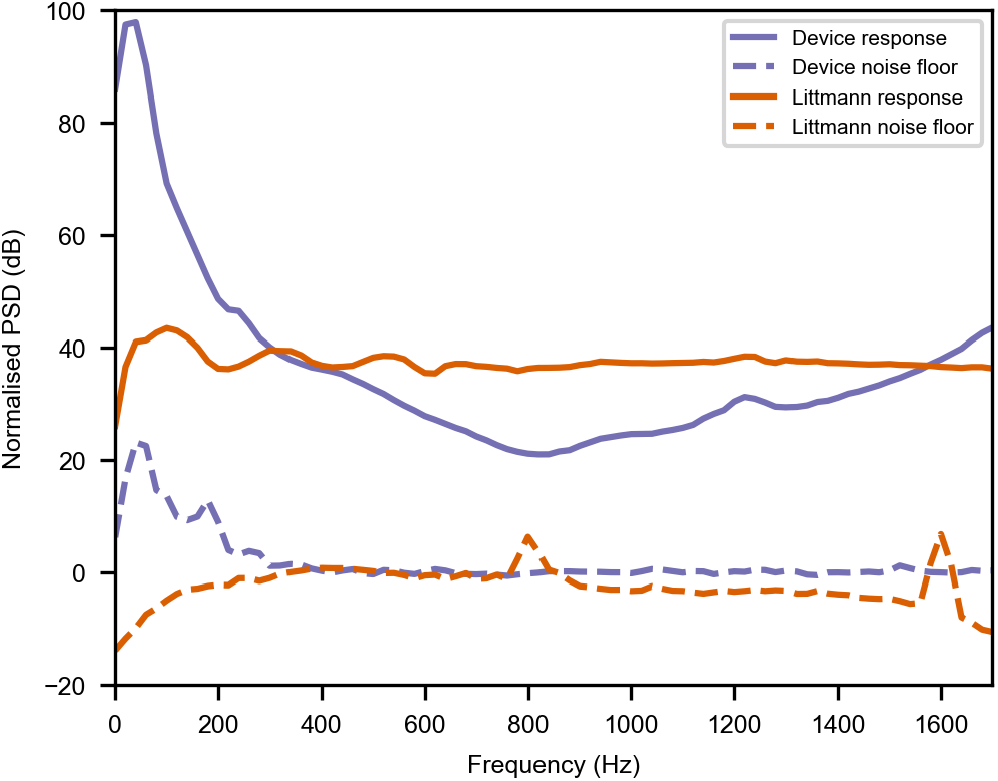
Response of a device sensor and a Littmann 3200 electronic stethoscope to the same excitation signal on a laboratory phantom. PSDs are referenced to the sensor’s noise floors (also shown). For details see section 4.2.

Our device sensor outperforms the Littmann 3200 at frequencies below 300 Hz, indicating enhanced sensitivity to the key frequency range of heart sounds. While the Littmann 3200 shows better performance above 300 Hz, the device sensor maintains a significant margin above its noisefloor. Although this paper focuses on the 20–200 Hz range, the validation demonstrates the sensor’s potential for detecting higher-frequency murmurs and respiratory sounds. Overall, the performance of both sensors is comparable, as expected, given their similar use of PZT discs for vibration detection.

### 2.3 Processing of heart vibration data

In a subject with a structurally normal heart, two major heart sounds, known as S1 and S2, are heard in rhythm [4]. The S1 sound, caused by the closure of the mitral and tricuspid valves at the start of the systolic period has a frequency range of 10-200 Hz [26], with power concentrated in the 25-45 Hz band [27]. The S2 sound is similarly caused by the closure of the aortic and pulmonary valves at the start of diastole and has a frequency range of 25-250 Hz [26] with power concentrated between 55-75 Hz [27]. The corresponding chest accelerations are of the order of milli-gravity (mg) [28] and have a duration of approximately 0.1 seconds [26]. The strength of S1 and S2 sounds across the chest is highly non-uniform, with S1 stronger towards the lower left of the chest, near the apex of the heart.

Figure 3a shows an example of the synchronised recordings obtained from a female subject at the aortic auscultation position at the upper left sternal border. Significant variations in signal strength can be observed across the multi-sensor device, illustrating the advantage of having a large sensing area. The position of the heart within the chest, combined with varying widths of fatty tissue and the ribs, leads to highly localised vibration zones on the chest. In addition, non-uniform application pressure from the hand and spurious movement can cause inter-sensor differences.

**Fig. 3.**
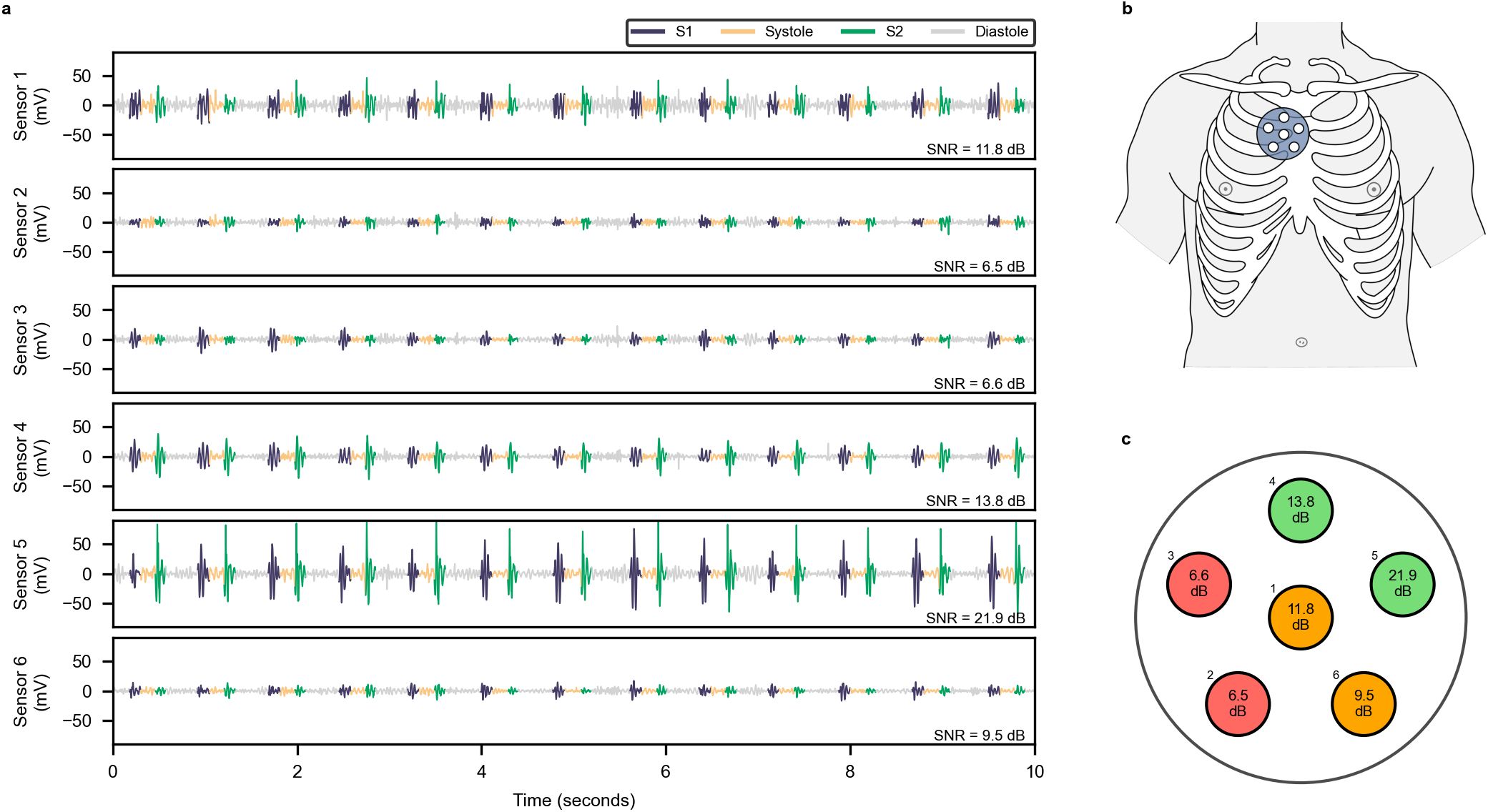
Example multi-sensor heart sound recording from the device, held by a female subject at the aortic auscultation site. **a**, Sensor outputs after a zero-phase 20 to 600 Hz bandpass filter. Significant variations in signal amplitude and SNR can be seen across the sensors, with sensor 5 achieving a clear peak of 21.9 dB. **b**, The aortic auscultation location on the chest, illustrating that the best recordings in this case were towards the sternal border. **c**, SNR results shown visually, with a traffic light system coding poor SNR (*<*8 dB), moderate SNR (8-12 dB), and good SNR (*>*12 dB).

Automated signal quality assessment algorithms are applied to the multi-sensor output to objectively select the best sensor for further analysis and decide if a recording needs to be repeated. We compute a signal-to-noise ratio (SNR) for each sensor in the recording, by comparing the power spectral density of S1 and S2 sounds to surrounding quiet systolic or diastolic sections. When used on the chest, sensors encounter background vibrations, contributing to mechanical noise. This noise, calculated as background noise in this paper, stems from room vibrations, biological processes in the body of the subject and the shaking of the user’s hand. These background vibration levels exceed the noise floor of the sensor shown in Figure 2. However, the high sensitivity of the sensors enables them to achieve high SNRs ensuring that sensor self-noise does not limit performance. The SNR algorithm is further described in Section 4.4.

Figure 3c shows the significant variation in SNR across the device. It also shows a potential way these SNR values could be displayed to a user to help guide optimal device placement. In this specific example, the user would need to reposition the device slightly northeast from their current position for better signal quality. A sensor signal with a high SNR (e.g. sensor 5 in Figure 3) would be selected for further analysis, either by a clinician (remotely or in-person) or a diagnostic machine learning model.

### 2.4 Validation study

A proof-of-concept study on healthy volunteers assessed the device’s performance at collecting heart sounds in the hands of untrained users. A total of 40 participants (20 male and 20 female), all without medical training, were recruited from Cambridge, UK. The participants represented a diverse range of ages and BMIs, as shown in Table 1. Participants were categorised into standard adult BMI categories of under- weight (BMI ≤ 18.5 kgm^−2^), normal (18.5 kgm^−2^ *<* BMI ≤ 25 kgm^−2^), overweight (25 kgm^−2^ *<* BMI ≤ 30 kgm^−2^), and obese (BMI *>* 30 kgm^−2^).

**Table 1.** Key biometric variables of study participants, split by biological sex.

| Variable | Group | Female (n=20) | Male (n=20) | p-value |
| --- | --- | --- | --- | --- |
| Age (years) |  | 45 (32-56) | 42 (28-56) | 0.61 |
| BMI ($\text{kgm}^{-2}$) | | 22.9 (21.1-28.5) | 26.6 (23.2-29.0) | 0.59 |
| BMI group | Underweight | 1 (5%) | 1 (5%) | - |
|  | Normal | 11 (55%) | 7 (35%) | - |
|  | Overweight | 4 (20%) | 7 (35%) | - |
|  | Obese | 4 (20%) | 5 (25%) | - |
| Chest size (cm)<sup>1</sup> |  | 90 (87-110) | 101 (95-109) | 0.28 |
| Bra | No bra | - | 20 (100%) | - |
|  | Bra without underwire | 6 (30%) | - | - |
|  | Bra with underwire | 14 (70%) | - | - |

Continuous variables are reported as median (interquartile range). Categorical variables are reported as frequency (proportion). Statistical difference between the means of the continuous variables was assessed using Welch’s *t*-test.
<sup>1</sup>Chest size was measured as circumference around the widest part of the chest.

Under supervision from a researcher, patients placed the patch at four different locations on their chests. Three of these are shown in Figure 1: the aortic area (second intercostal space [ICS], right sternal border), tricuspid area (fourth ICS, left sternal border), and mitral area (here defined as fifth ICS, midclavicular line). The mitral location is typically found by palpating for the apex beat. However, to make this recording accessible for untrained users, the midclavicular line was used as an anatomical reference point. Additionally, a nearby ‘mitral left’ location was included in the protocol, where the patch was moved left (from the patient perspective) from the mitral site so that the patch edge was located on the midclavicular line. This ensured comprehensive coverage of the mitral region. Extended Data Figure E2 details these four locations. At each location, three 15-second recordings were made over a thin layer of clothing such as a T-shirt or blouse. For participants wearing a bra, recordings were made both with and without it to investigate the effect of the bra on recording quality. See Methods 4.3 for further details on the study protocol.

All recordings were segmented using the machine learning algorithm from Section 2.3. A researcher manually checked all segmentations. Recordings where the researcher could not confidently validate the segmentation due to excessive noise or low signal strength were labelled as ‘unsegmentable’. An SNR was then calculated for each of the six sensors in every segmentable recording. The sensor with the highest SNR within any 10-second window was selected as the ‘best’ SNR for that recording. This process avoided unwanted noise in certain brief sections of the recording (e.g. start or end). A recording was deemed to ‘pass’ the signal quality assessment if it met the following criteria: (i) a valid segmentation, (ii) at least six complete sections of S1, S2, systole, and diastole, and (iii) a sensor with an SNR above a chosen threshold (8 dB in this paper, as explained below). Table 2 presents the pass rate for patients across all recording locations, categorised by sex and BMI. For males, performance was excellent at the three main auscultation sites (aortic, mitral, tricuspid), with every male participant achieving a passable recording at the tricuspid and mitral sites, regardless of BMI. For normal or underweight female participants, similar high performance was observed. However, the performance was reduced for females with higher BMIs at the tricuspid and mitral sites. Specifically, 75% of overweight or obese patients had a successful tricuspid recording, while only 50% of obese patients achieved a passable mitral recording. The impact of BMI was further compounded when recordings were attempted whilst patients wore a bra. Mitral recordings with a bra were successful in 75% (n=3) of overweight participants but only 25% (n=1) of obese participants.

**Table 2.** Per-location signal quality assessment pass rate for patients with different sex and BMIs.

| Recording location | Signal quality pass rate (%) |  |  |  |  |  |
| --- | --- | --- | --- | --- | --- | --- |
|  | Female BMI |  |  | Male BMI |  |  |
|  | < 25<br>(n=12) | 25 – 30<br>(n=4) | > 30<br>(n=4) | < 25<br>(n=8) | 25 – 30<br>(n=8) | > 30<br>(n=4) |
| Aortic | 100 | 100 | 100 | 88 | 88 | 100 |
| Tricuspid | 92 | 75 | 75 | 100 | 100 | 100 |
| Mitral | 92 | 100 | 50 | 100 | 100 | 100 |
| Mitral Left | 100 | 75 | 75 | 100 | 62 | 75 |
| Mitral (with bra) | 83 | 75 | 25 |  |  |  |
| Mitral Left (with bra) | 92 | 25 | 50 |  |  |  |

All BMI values are in $\text{kgm}^{-2}$ . A recording location passed for a patient if any of the three repeats passed.

Most participants (n=32) achieved a passable recording at both the mitral and ‘mitral left’ recording sites. Two participants had a passable recording at the ‘mitral left’ location alone, whereas five participants had a passable recording at only the mitral location. Only one participant failed to get a passable recording at either of the mitral sites.

The critical importance of using multiple sensors over a single-sensor device was systematically examined. Table 3 shows that the sensor with the highest SNR varied significantly across different auscultation sites. For the aortic site, sensors 5 and 6, positioned near the sternum (see Figure 3c), consistently provided the best recordings. Notably, the central sensor, which users aimed to align with the recording site, was never the optimal choice. Table 3 also shows that the pass rate across different locations would have considerably decreased if only the central sensor in the device was available. This reduction in performance was particularly pronounced in overweight and obese patients. For instance, at the aortic site the pass rate for these patients was 83% using the best of 6 sensors, compared to just 32% when relying solely on the central sensor.

**Table 3.** Advantages of a multiple sensor device, broken down by recording location. The first part of the table shows the frequency with which each sensor had the best SNR. The second part shows how the signal quality pass rate changes between using the best of 6 sensors, or just the central sensor in the device.

|  | Frequency |  |  |  |  |  | Pass rate (%) |  |  |  |
| --- | --- | --- | --- | --- | --- | --- | --- | --- | --- | --- |
|  | best sensor |  |  |  |  |  | BMI<25 |  | BMI>25 |  |
|  | 1 | 2 | 3 | 4 | 5 | 6 | Best | Central | Best | Central |
| Aortic | 5 | 1 | 1 | 9 | **55** | 39 | 90 | 60 | 83 | 32 |
| Tricuspid | 22 | 14 | **25** | 9 | 18 | **25** | 93 | 88 | 85 | 60 |
| Mitral | 20 | 7 | 27 | **39** | 6 | 8 | 90 | 85 | 72 | 50 |
| Mitral Left | 13 | 12 | **44** | 22 | 3 | 1 | 87 | 70 | 53 | 28 |
| Mitral (Bra) | 4 | 3 | **13** | **13** | 0 | 6 | 81 | 72 | 33 | 17 |
| Mitral Left (Bra) | 5 | 3 | 13 | **14** | 4 | 4 | 86 | 61 | 33 | 8 |

Patients made three recordings at each site - each recording repeat is reported separately. The best sensors at each location are shown in bold.

These pass rates for the recordings are dependent on the SNR threshold. The value of 8 dB was selected to ensure clear S1 or S2 sounds in the recording and a significant gap between signal and noise for future usage on patients with abnormal murmurs that may have lower signal power than the major heart sounds. However, other thresholds could be picked depending on the exact clinical usage. Extended Fig. E3 shows the distribution of SNRs for all segmentable recordings in the dataset, and shows how the pass rate varies with the choice of threshold. The use of multiple sensors significantly improves the recorded SNR compared to a single sensor, regardless of the chosen threshold.

Beyond a binary pass criterion, the effect of recording location, sex, and BMI on the SNR distribution was further investigated, as shown in Figure 4. Device performance was highest at the tricuspid site, where 73% of recordings had an SNR greater than 12 dB. However, when comparing male and female participants (Figure 4b), SNR at the tricuspid site was significantly lower for female patients (*p <* 0.001). Figure 4b also indicates that female SNRs were higher than males at the aortic (*p* = 0.016) and mitral left (*p* = 0.036) sites. However, BMI is a significant confounder in these results, with no statistically significant differences at these sites among non-overweight patients. In fact, Figure 4c shows that overweight or obese participants have significantly lower SNRs (*p <* 0.001) at all locations except the tricuspid site.

**Fig. 4.**
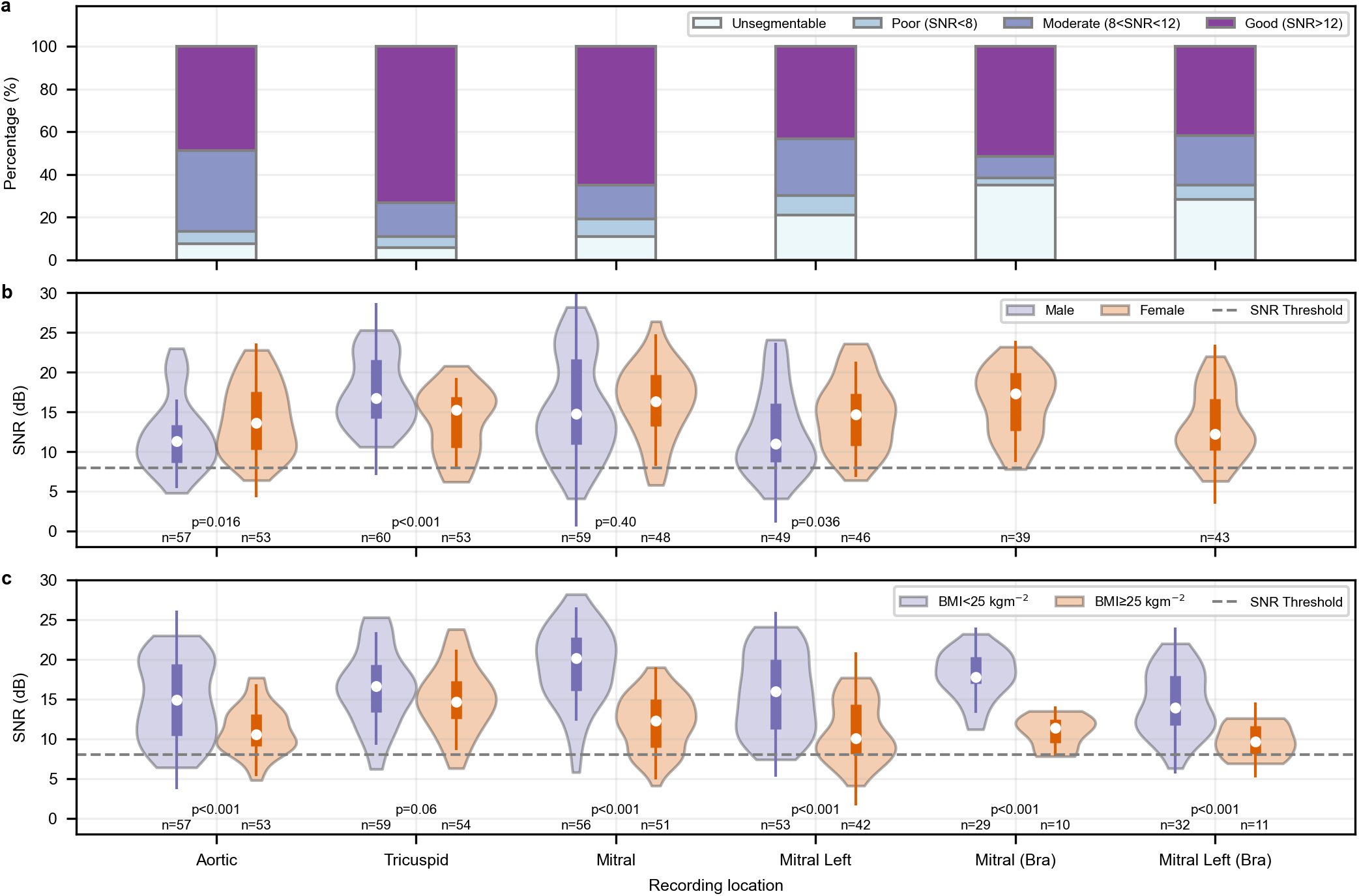
Distribution of recording signal-to-noise ratios (SNRs) by key variables. **a**, Proportion of unsegmentable and segmentable recordings for each chest location, with segmentable recordings categorised into poor, moderate, or good SNR ranges. **b**, SNR of segmentable recordings, split by biological sex. **c**, SNR of segmentable recordings, split into normal and high (overweight or obese) BMI. The means of the distributions were compared using Welch’s t-test, with p-values shown.

## 3 Discussion

This paper presents a novel medical device designed to simplify the process of obtaining high-quality heart sounds, particularly for untrained users. Notably, the ability to use the device over clothing could significantly improve both comfort and adoption rates, especially for female participants in routine screening programs. The device’s large, flexible sensing area significantly improves the quality of signals recorded compared to a single central sensor. The objective signal quality assessment we developed provides useful feedback, guiding users as they position the device over key auscultation sites.

The current design is in its alpha prototype phase, and future work will focus on progressing towards medical device certification. This will require the design of a manufacturing process, but the sequential bottom-up construction of the device means techniques such as silicone overmoulding may be used. A key advantage of our design is that all the device components can be easily available at scale, requiring no specialised manufacturing processes.

The prototype currently uses a wired connection to an external data acquisition unit, but transitioning to a fully wireless device would enhance usability. This could be achieved by integrating a simultaneous ADC and Bluetooth Low Energy chip into the existing circuitry. The primary challenge in this transition will be maintaining the device flexibility while accommodating the additional components.

The participant study described in this paper serves as an initial validation of the device’s use by untrained users, particularly its functionality over clothing. The procedure was an example of a potential use-case of the device, where a trained professional is present in the room to supervise the use of the device but does not make physical contact with the patient. This setup simulates a common telemedicine usecase, where a clinician may supervise the process remotely without direct contact. The device has particular value in screening communities often missed by primary care, where patients may prefer minimal physical interaction or remain fully clothed. However, further studies are needed to assess its usability by unsupervised patients, which will require the development of straightforward instructional materials.

In our study, the device performed well for male participants and for underweight or normal-weight females, with and without a bra. However, a decrease in performance was observed for overweight and obese female participants, particularly at the mitral site, likely due to the attenuation of heart vibrations by additional tissue. The presence of a bra likely further contributes to this effect. A couple of female participants also advised that they had breast implants, which reduced the signal quality further at the tricuspid site, likely due to the added damping from the implant material. Future iterations of the device would integrate real-time signal quality feedback, guiding users on the direction they should move the patch to obtain an optimum signal, particularly in challenging cases. Female participants could be asked to first attempt recordings with a bra, and then remove it if the signal quality is insufficient.

A limitation of the validation study is the small sample size, particularly for sub-analyses of the effect of BMI and wearing a bra. A second limitation is the absence of participants with cardiovascular disease. However, laboratory testing demonstrates that the device should be capable of detecting higher-frequency murmurs, and the signal quality assessment is designed to be robust in the presence of systolic or diastolic murmurs. Although the volunteer pool included a range of ages, as a screening device, the target population will predominantly be older, more likely to have hand tremors and less familiar with apps and technology. The device will be prospectively tested on a more representative population in a future clinical study, where it is combined with a diagnostic machine learning algorithm [24] to provide a full screening solution for valvular heart disease.

## 4 Methods

### 4.1 Device construction

The substrate of the device consists of a 4 mm thick layer of highly-damped silicone gel (Ecoflex™ gel with Silc Pig™ white pigment), which is cast in a mould. The base of the substrate is covered with a very thin layer of polyethylene to provide insulation and ingress protection. The individual sensors are mounted onto the polyethylene base layer during assembly (using double-sided adhesive tape), and the silicone gel is then moulded around them. Two layers of acoustic foam above the gel substrate provide protection to the device internals and provide minor additional rigidity so that the device can be easily held in one hand. The layers of foam are not glued in place, and there is air between them and the surrounding layers, increasing the flexibility of the device. Finally, a silicone top layer is bonded to the substrate to yield a fully encased, waterproof device that is safe for patient contact.

Each vibration sensor in the device consists of a lead zirconate titanate (PZT) disc transducer housed in a circular 3D-printed casing with an outer diameter of 18 mm. The PZT discs are rim-mounted to the support structure via a foam layer that ensures contact with the chest when the sensor is pressed against it. When a vibration sensor is pressed against the chest, it forms a coupled system with the human body, where the operator’s hand not only provides mechanical impedance but also affects the properties of the chest tissue and the skin surface topology. MEMS accelerometers, air-coupled electret microphones, and piezoelectric contact microphones are commonly used to capture body sounds. However, MEMS accelerometers exhibit reduced performance when held, as the grip impedes their motion. In contrast, pressure sensors perform optimally when held, as the firm contact maximises the pressure applied to the sensor, similar to the mechanism of traditional acoustic stethoscopes [29]. Among these, piezoelectric contact microphones are preferable for this application due to their reduced sensitivity to airborne noise and manufacturing variations, compared to air-coupled sensors.

Each device sensor is connected to three wires: a reference voltage, a supply voltage, and signal output channel. A shielded cable is used to connect the device to a ‘hub’ circuit board fitted to two NIc9232 input modules mounted on an NI 9174 compact data acquisition chassis. The ‘hub’ circuit board is used to provide a power source for the operational amplifiers in the form of three 1.5 V AA batteries. The six sensors on the device are sampled simultaneously at a sampling frequency of 5,120 Hz, with anti-aliasing filtering provided by the data acquisition unit.

### 4.2 Laboratory testing

To characterise a sensor for use on the human chest the typical ratio between the heart sound signals it can pick up and its own noise floor must be established. However, the level of excitation on the human chest can display significant inter- and intrasubject variation. For a more repeatable comparison between sensors, we employ a simple laboratory setup (“phantom”) consisting of a cylinder of silicone elastomer (Ecoflex 00-10), excited by an electrodynamic shaker.

To validate the sensors used in this device, we compare their performance to that of a Littmann 3200 electronic stethoscope, a device which is often used to benchmark new heart sound sensors.

First, we establish an estimate for the noisefloor for each sensor. This is done by hanging the sensor in air in a quiet room. We take a 10 second segment for the middle of a longer recording (to counteract and edge effects) and compute the PSD using Welch’s method with Hann windows, 50% overlap and a 20 Hz frequency resolution. The measured noisefloors of a Littmann 3200 and one of the sensors in the device are shown in Figure 2. For the Littmann 3200 bandpass filtering effects are clearly evident. For both sensors the noisefloor between 500 and 1000 Hz is fairly constant (neglecting a spike at 800 Hz for the Littmann 3200), and we take the mean PSD in this frequency range as a reference level. We note that for the device sensor the noisefloor above 300 Hz is set by the noise level on the NI data acquisition unit, rather than by the sensor itself.

Next, we measure the response of both sensors to the same excitation signal on our phantom. The excitation signal can be controlled to yield a desired output level, and here we use a signal designed to give a fairly uniform response for the Littmann 3200 between 20 and 1800 Hz, at a mean level of 36 dB above the reference noise level. At frequencies below 200 Hz the amplitude of normal heart sounds (male, normal BMI) typically exceeds this amplitude, while above 200 Hz the amplitude of normal heart sounds is typically lower. We have extended the excitation to 1800 Hz in order to establish the suitability of the sensors for detecting murmurs and other higher frequency body sounds.

Figure 2 shows the PSD levels measured on the phantom, normalised by the reference noise level for each sensor. This allows us to compare the response amplitudes, and also assess whether body sounds at the chosen amplitude level could be distinguished from the noisefloor of the sensors.

### 4.3 Participant study

The adult participants (n=40) were invited through departmental bulletins within the university, or direct contact by a researcher. Participants were excluded if they had a chest wound, any known cardiovascular disease, or had received any prior clinical training. All participants provided informed, written consent. The study was approved by the Ethics Committee of the Department of Engineering, University of Cambridge under code 489.

To ensure the study protocol was correctly followed, an expert researcher was in the room to ensure the device was placed at approximately the correct location on the chest. Participants were asked to remove outer layers so that they were just wearing a T-shirt, blouse, or similar. The researcher briefly instructed the patient on how to place the patch by feeling for their intercostal spaces or other anatomical features as appropriate. Participants were then asked to place the centre of the patch at the identified location and apply moderate, uniform pressure with their palm.

Standard biometric data (age, sex, weight, height and chest circumference) was recorded for each patient. At the end of the session, participants were asked to complete a short questionnaire on how they found the usability of the device and the recording process.

### 4.4 Quantitative signal quality assessment

We define a signal-to-noise ratio (SNR) for heart sound signals, following previous work [14], to objectively measure the clarity of the S1 and S2 sounds relative to the silent sections of the recording. Previous methods relied on peak-to-peak time series measurements [14], which are highly variable across devices due to different digital filters affecting signal morphology. In this work, we compute an SNR based on a comparison of the power spectral density (PSD) of the signal sounds in a fixed frequency band.

The full SNR computation method, shown in Fig 5, requires that the signal first be segmented into S1, systole, S2, and diastole sections (Fig 5a). We use a recurrent neural network and hidden semi-Markov model algorithm to automate this process [24], which previously won the George B. Moody PhysioNet Challenge 2022 [11]. Any segmented sections with high amplitude spikes (e.g. due to stethoscope movement) are removed, by searching for any sections where the maximum amplitude of the section is more than three times the 98% quantile for all sections of that heart sound state.

**Fig. 5.**
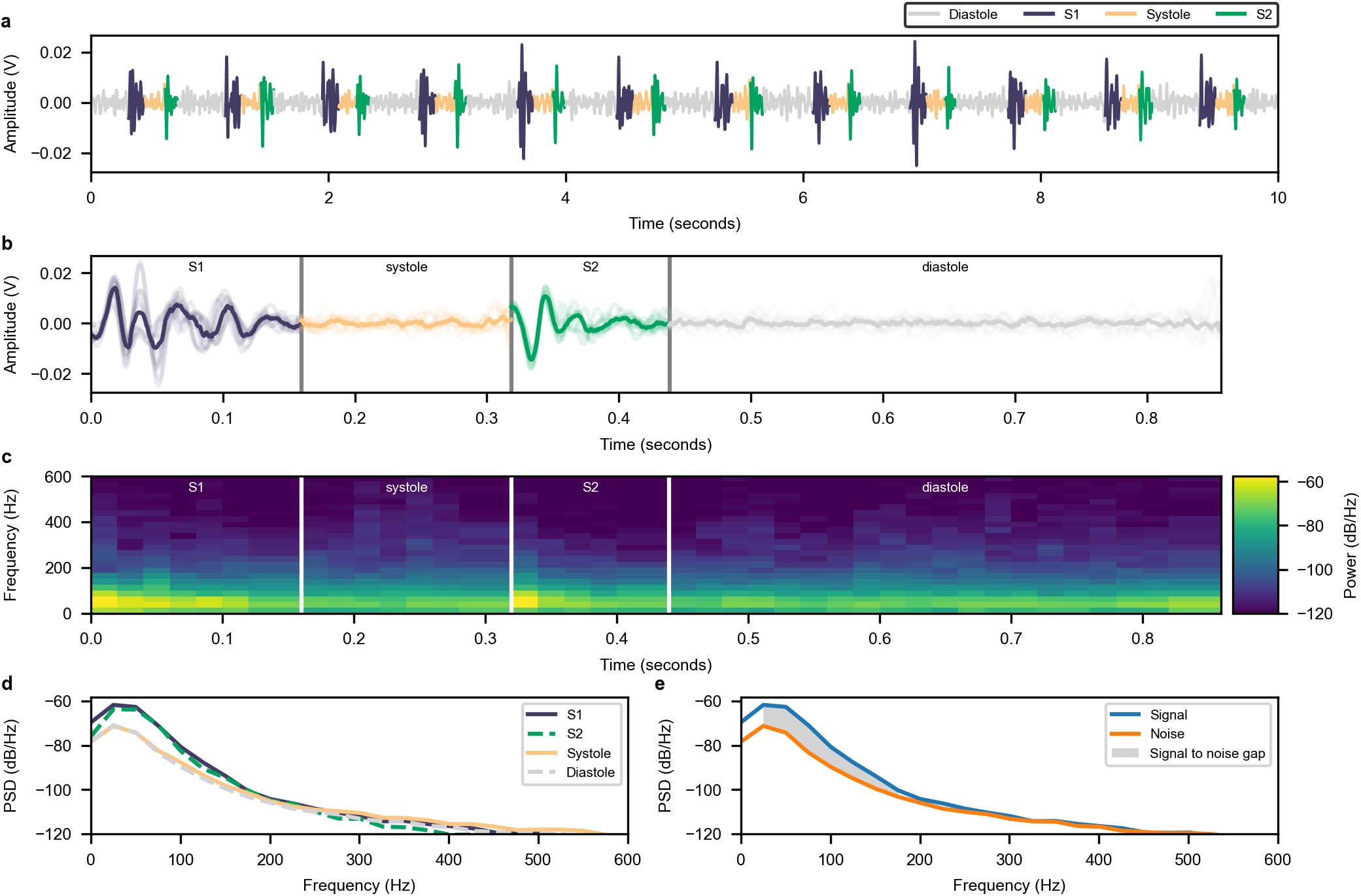
Computation of signal-to-noise ratio (SNR) for a moderate-quality heart sound signal. **a**, A heart sound signal segmented using recurrent neural network algorithm. **b**, Individual heart sound states are aligned using their cross-correlation. Shown is each individual state section and their median in bold. **c**, Median spectrogram is computed of the aligned heart sound states. **d**, Power spectral density (PSD) is computed for each state by averaging the power in each spectrogram frame, with 1*V* ^2^*/*Hz as a reference. **e**, The signal-to-noise gap is taken as the maximum sound PSD (S1, S2) compared to the silent state PSDs (systole, diastole). The final SNR calculated is 9.3 dB, indicating a moderate-quality signal with some background noise.

Next the individual S1 and S2 sounds are aligned using their cross-correlation to allow for slight time offsets in their segmentation. All states are then padded or truncated to the lower quartile of their durations (Fig 5b).

We then compute a spectrogram for each heart sound state section, and take a median to form a median spectrogram representation of the heartbeat (Fig 5c). The median is used to discard spectrogram frames that have large energies due to recording spikes and keep only time-frequency content that repeats with every heartbeat. A Hann window is used with a length of 40 ms and step of 20 ms. This gives a frequency resolution of 25 Hz. A power spectral density for each heart sound state is then calculated by taking the mean of each spectrogram section over time (Fig 5d). To create the ‘signal’ PSD, we then take the maximum of the S1 or S2 PSDs in each frequency bin. To create the ‘noise’ PSD, we take the minimum of the systole or diastole PSDs across frequency (Fig 5e). The SNR, as a function of frequency, is then calculated by simply dividing the signal by the noise PSD in V^2^*/*Hz. To produce a single value, we take the mean of the SNR in the 25 ≤ *f <* 200 Hz range, which contains the majority of S1 and S2 energies [26]. We finally report values in decibels by transforming to a logarithmic scale:

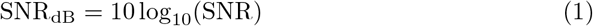

In contrast to previous approaches, this SNR is invariant to digital filtering of the signal, as the filtering impacts both the noise and signal equally. As a result, the effects cancel out when their ratio is computed.

The method described does not consider the presence of murmur sounds, as only healthy patients were included in the validation study. However, it could trivially be adapted for abnormal sounds, by including the segmented ‘murmur’ state as a ‘signal’ PSD and extending the frequency range beyond 200 Hz.

## Data Availability

All data produced in the present study are available upon reasonable request to the authors.

## Acknowledgements

We acknowledge the support of the UK Medical Research Council (MRC) through its Confidence in Concept grant, the Engineering and Physical Sciences Research Council (EPSRC) through its Impact Accelerator Award, and Emmanuel College, Cambridge. We acknowledge John Hazlewood for his assistance in the acoustics lab, Anthony Luckett for the 3D printing of sensor casings and Catia Santos, Paul Pedersen and especially Mark Huntsman for production and assembly of the PCBs and wiring for the device described in this paper.

## Author contributions

MN, AM, and AA designed the device prototype with clinical input from RS. MN performed sensor benchmarking. AM, MN, and AA developed the signal processing algorithms and data analysis. MN, AM, and NR designed and performed the human subject study with supervision from RS and AA. AM, MN, and AA drafted the manuscript. All authors reviewed and contributed to the final version.

## Competing interests

MN, AM, and AA are authors on a patent application related to the device design in this work (PCT/GB2024/051172).

## Appendix A Extended Data

**Extended Fig. E1.**
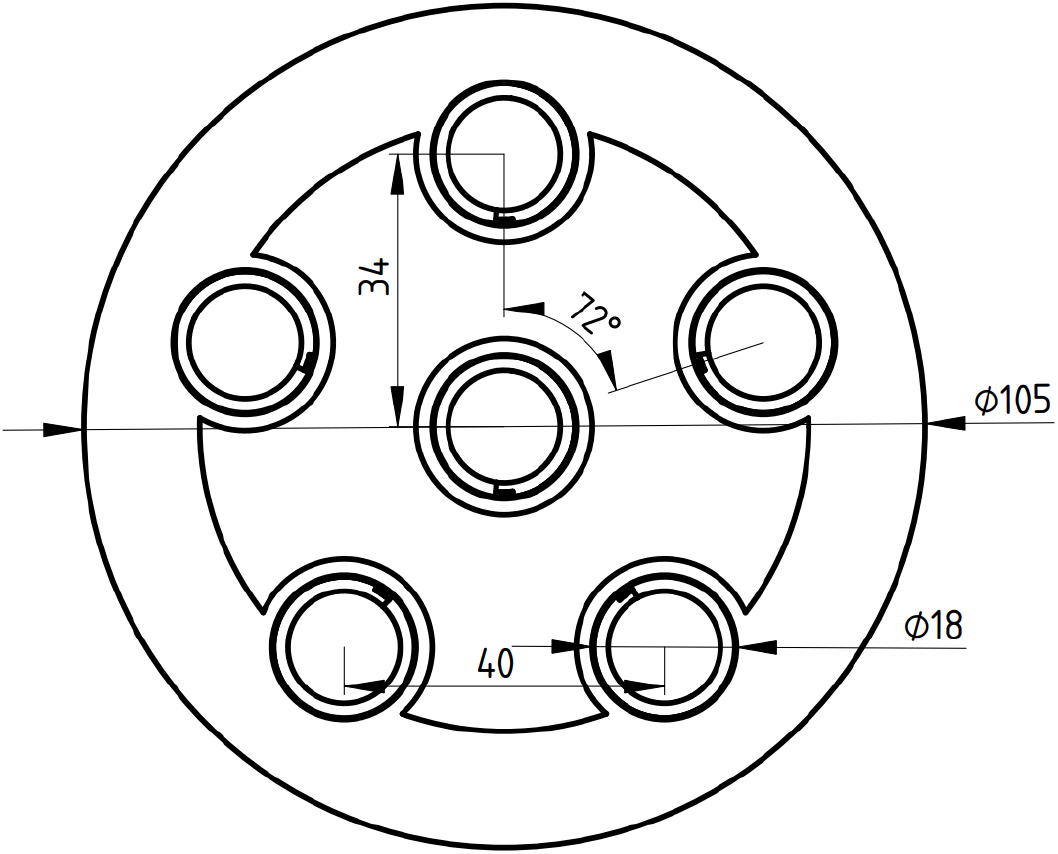
2D layout of sensors within the device. Sensors are arranged in a pentagon, with a single sensor in the centre. All dimensions are in millimetres.

**Extended Fig. E2.**
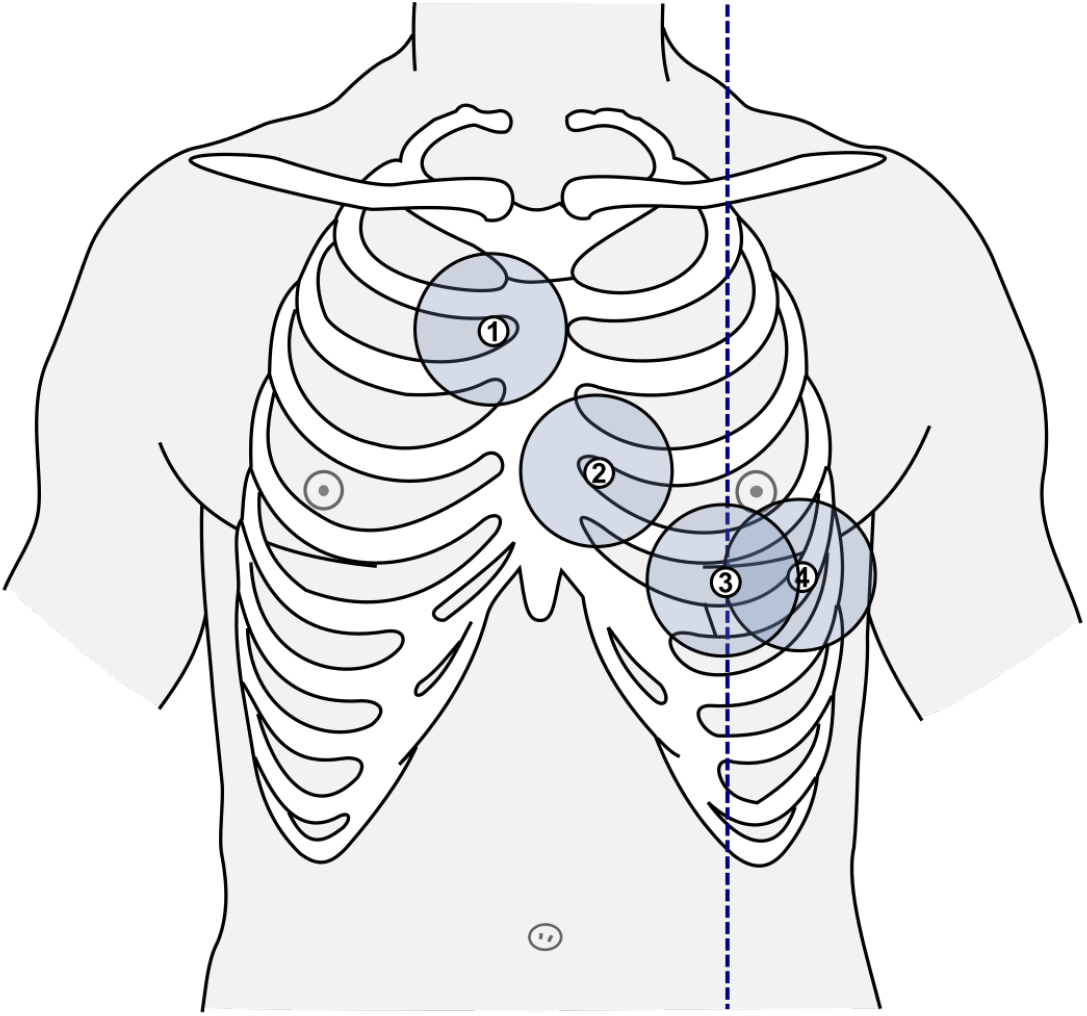
Locations recorded by patients. (1): Aortic area, second intercostal space (ICS) on the right sternal border. (2): Tricuspid area, fourth ICS on left sternal border. (3): Mitral area, under left breast (approximately fifth ICS) on midclavicular line (dashed blue line). (4): Mitral left area, under left breast with patch edge on midclavicular line.

**Extended Fig. E3.**
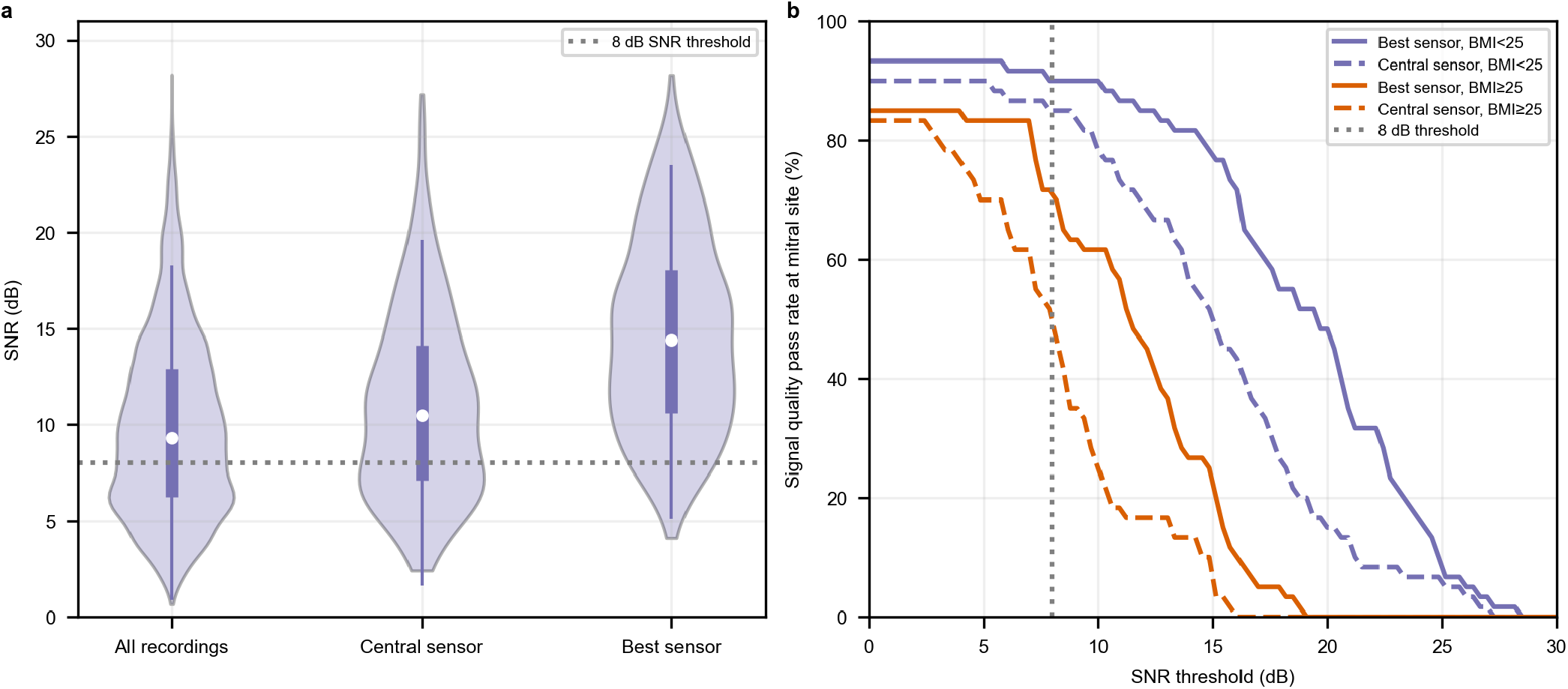
Sensitivity analysis of pass rate on SNR threshold. (a) shows the SNR distribution for segmentable recordings, for all sensors in the dataset, just the central sensor, and the chosen best sensor. (b) shows how the signal quality pass rate at the mitral site varies depending on the SNR threshold chosen. This is split into non-overweight and obese/overweight patients.

## Notes

### Funding Statement

The study was supported by the UK Medical Research Council (MRC) through its Confidence in Concept grant, the Engineering and Physical Sciences Research Council (EPSRC) through its Impact Accelerator Award, and Emmanuel College, Cambridge.

### Author Declarations

The Ethics Committee of the Department of Engineering, University of Cambridge gave ethical approval for this work (project code 489).

